# A multi-phenotype approach implicates *SH2B3* in the genetics of chronic kidney disease

**DOI:** 10.1101/2024.06.29.24309709

**Authors:** Kim N. Tran, Heidi G. Sutherland, Andrew J. Mallett, Lyn R. Griffiths, Rodney A. Lea

## Abstract

Chronic kidney disease (CKD) is a complex condition with diverse underlying causes that lead to a progressive decline in kidney function. Genome-wide association studies (GWASs) have identified numerous genetic loci associated with CKD, yet much of the genetic basis remains unexplained. Part of the reason is that most GWASs have only assessed kidney function via single biomarkers such as estimated glomerular filtration rate (eGFR). This study employs a novel multi-phenotype approach, combinatorial Principal Component Analysis (cPCA), to better understand the genetic architecture of CKD. Utilizing a discovery cohort of white British individuals from the UK Biobank (n=337,112), we analyzed 21 CKD-related phenotypes using cPCA to generate over 2 million composite phenotypes (CPs). More than 46,000 CPs demonstrated superior performance in classifying clinical CKD compared to any single biomarker, and those CPs were most frequently comprised of eGFR, cystatin C, HbA1c, microalbuminuria, albumin, and LDL. GWASs of the top 1,000 CPs revealed seven novel genetic loci, with *CST3* and *SH2B3* successfully replicated in an independent Irish cohort (n=11,106). Notably, the index SNP of the *SH2B3* gene, which encodes a regulator in immune responses and cytokine signaling, is a loss-of-function variant with a combined beta of −0.046 and a p-value of 3.1E-56. These results highlight the effectiveness of a multi-phenotype approach in GWASs and implicate a novel functional variant in SH2B3 in CKD phenotypes.

**TRANSLATIONAL STATEMENT:** The application of combinatorial Principal Component Analysis (cPCA) in our study has identified *SH2B3* as a novel genetic locus associated with chronic kidney disease (CKD). This discovery advances our understanding of CKD’s genetic architecture beyond single biomarker analyses, potentially leading to more precise diagnostic tools and personalized treatment strategies. Future research should focus on validating these findings in diverse populations and integrating cPCA-derived biomarkers into clinical practice to enhance CKD prediction and management, ultimately improving patient outcomes.

## INTRODUCTION

Chronic kidney disease (CKD) is a collective term encompassing a range of heterogeneous diseases characterized by persistent structural or functional kidney abnormalities. CKD is stratified into five stages, culminating in kidney failure, which necessitates consideration of interventions such as kidney transplantation or dialysis. This condition has a high prevalence, affecting approximately 10-15% of the global population, resulting in significant burden on both public health and the economy.^1^

Genome-wide association studies (GWAS) investigating CKD have traditionally focused on evaluating kidney function using single biomarkers, such as estimated glomerular filtration rate (eGFR), microalbuminuria, or blood urea nitrogen.^2–5^ For example, a robust GWAS analysis of eGFR in a cohort of over 1.2 million individuals identified 634 independent genetic signals, collectively accounting for 9.8% of the eGFR variance.^4^ However, a portion of the heritability of CKD remains unexplained. This gap in understanding can be attributed, in part, to the fact that eGFR and other individual biomarkers do not fully capture the underlying causes of CKD nor accurately predict an individual’s risk of CKD or progression to kidney failure.^6^ For a comprehensive diagnosis and prognosis of systemic CKD, it is recommended to employ a combination of various markers that collectively reflect the diverse alterations occurring during the course of CKD.^7^

Previously, we employed principal component analysis (PCA) on multiple quantitative phenotypes associated with CKD, uncovering a novel susceptibility gene for kidney function that remained undetected in single-phenotype GWASs.^8^ In this study, we introduce and implement a new approach termed combinatorial PCA to further investigate the genetic basis of CKD within the UK Biobank dataset. As a result, we identified a new locus, *SH2B3* (SH2B adaptor protein 3), to be associated with CKD.

## METHODS

### Research cohort

The UK Biobank (UKB) is a longitudinal cohort study examining the interplay between genes, the environment, and health. It encompasses over 500,000 participants aged 40-69 years, recruited between 2006 and 2010 from 22 assessment centers across England, Scotland, and Wales. Approval for the UKB study was obtained from the North West Multi-Centre Research Ethics Committee, and all participants provided written informed consent. This research has been conducted utilizing the UK Biobank Resource under Application Number 60111.

We selected White-British samples, constituting the largest ethnic group within the UKB dataset, for the discovery cohort based on both the ‘Ethnic background’ and ‘Genetic ethnic grouping’ data. This approach allowed us to accurately identify individuals who self-identified as ‘White British’ and exhibited very similar genetic ancestry profiles, as determined by a principal components analysis of their genotypic data. Additionally, we excluded individuals whose genetic sex differed from their self-identified sex, those with sex chromosome aneuploidy, or those who were not included in the genetic principal components analysis conducted by the UKB research team. The final sample size was 337,112. Finally, we included individuals from the Irish ethnicity within the UKB dataset for the replication analyses (n=11,106). The data processing steps were performed similarly to those used for the discovery cohort.

### Phenotype data

In total, 21 biomarkers relevant to chronic kidney disease (CKD) were included in this study (Table 1). These phenotypes were assessed based on the correlations of the measurements with prevalence of CKD, CKD stages, kidney function, and an increased risk of adverse outcomes in individuals with CKD. All measurements were collected at baseline for all participants. Details of the assay manufacturers, analytical platforms, and analysis methodologies can be found at https://www.ukbiobank.ac.uk/enable-your-research/about-our-data/biomarker-data. Quantitative measures outside their respective analytical ranges were treated as missing data. Estimated GFR (eGFR) was calculated using the creatinine-based CKD-EPI-2021 equation without race coefficient.^9^ Samples with more than 30% missing data points were excluded. Remaining missing phenotypic values were imputed to obtain a complete dataset using the R package missMDA v1.11,^10^ ensuring that the imputed values had no effect on the principal component analysis (PCA) results. PCA was performed using the R package FactoMineR v1.34.^11^

**Table 1.** 21 kidney function related phenotypes selected from the UK Biobank dataset.

| No. | Phenotypes | Abbr. | Relation to kidney function | References |
| --- | --- | --- | --- | --- |
| 1 | Albumin | ALB | Associated with reduced kidney functions in HIV-infected individuals and elders | Lang et al<sup>12,13</sup> |
| 2 | Apolipoprotein A | APOA1 | Higher APOA1 associated with lower CKD prevalence | Goek et al<sup>14</sup> |
| 3 | Apolipoprotein B | APOB | Higher APOB associated with lower eGFR, increased ESRD risk | Zhao et al<sup>15</sup>, Zhao et al<sup>16</sup>, Kwon et al<sup>17</sup> |
| 4 | Body mass index | BMI | Higher BMI associated with increased risk of CKD | Ejerblad et al<sup>18</sup>, Lu et al<sup>19</sup>, Herrington et al<sup>20</sup> |
| 5 | Calcium | CALC | Lower CALC associated with rapid CKD progression | Janmaat et al<sup>21</sup> |
| 6 | C-reactive protein | CRP | Higher CRP associated with CKD incidence | Fox et al<sup>22</sup> |
| 7 | Cystatin C | CYSC | CYSC levels associated with kidney function | Benoit et al<sup>23</sup> |
| 8 | Diastolic blood pressure | DBP | Lower DBP associated with increased mortality in CKD patients | Agarwal<sup>24</sup>, Mitka<sup>25</sup> |
| 9 | Creatinine-based eGFR (CKD-EPI Creatinine Equation - 2021) | eGFR | Marker of kidney function | Inker et al<sup>9</sup> |
| 10 | Gamma glutamyltransferase | GGT | Higher GGT associated with increased risk of ESRD | Lee et al<sup>26</sup> |
| 11 | Glycated haemoglobin | HbA1c | Higher HbA1c associated with increased risk of CKD or CVD | Hernandez et al<sup>27</sup> |
| 12 | Haematocrit percentage | HCT | Lower HCT associated with declined kidney function and increased risk of ESRD | Iseki et al<sup>28</sup>, Chen et al<sup>29</sup> |
| 13 | HDL cholesterol | HDL | Both low and high HDL associated with adverse outcomes in patients with CKD | Nam et al<sup>30</sup> |
| 14 | LDL direct | LDL | Higher LDL associated with increased risk of CVD in non-dialysis CKD patients | De Nicola et al<sup>31</sup> |
| 15 | Microalbuminuria | MA | Biomarker for kidney injury | Glasscock<sup>32</sup> |
| 16 | Phosphate | PHOS | High PHOS associated with increased CVD risk and mortality in patients with or without CKD | Vervloet et al<sup>33</sup> |
| 17 | Systolic blood pressure | SBP | Lower SBP associated with ESRD and increased mortality in CKD patients | Agarwal<sup>24</sup> |
| 18 | Triglycerides | TRIG | Associated with CKD stages | Zubovic et al<sup>34</sup> |
| 19 | Urate | UA | Higher UA associated with new and progressive CKD | Oluwo and Scialla<sup>35</sup> |
| 20 | Urea | UREA | Higher UREA levels associated with adverse renal outcomes | Seki et al<sup>36</sup> |
| 21 | Vitamin D | VITD | Lower VITD associated with adverse outcomes and mortality in CKD patients | Kim and Kim<sup>37</sup> |

### Genotype data

Genome-wide genotyping was conducted on all UKB participants using the UK Biobank Axiom Array. Approximately 850,000 variants were directly measured, while over 90 million variants were imputed using the Haplotype Reference Consortium and UK10K + 1000 Genomes reference panels. Imputation data were stored in the compressed and indexed BGENv1.2 format. We converted the data from BGEN format into binary PGEN files and performed quality control procedures within PLINK2.0.^38^ The criteria for selecting variants were: (1) autosomal variants; (2) missing rate of less than 5%; (3) not significantly deviated from Hardy-Weinberg equilibrium (p-value=10E-10); (4) minor allele frequency (MAF) of at least 0.001; and (5) imputation score of more than 0.8. After quality control, we retained 12.7 million SNPs for subsequent analysis.

### CKD clinical outcome data

Health-related outcome data are available in death, hospital, and primary care records. Using the ICD-10 and ICD-9 codes (International Classification of Diseases, tenth and ninth editions), we categorized individuals diagnosed with chronic kidney disease, renal failure, renal sclerosis, chronic glomerulonephritis, nephritis, nephropathy, hypertensive chronic kidney disease, hypertensive heart and kidney disease, diabetes with renal complications, kidney replaced by transplant, disorders resulting from impaired renal function, or unspecified disorders of the kidney and ureter as CKD cases.

### Combinatorial principal component analysis (cPCA)

#### Principles

We developed an approach called combinatorial PCA (cPCA) to identify combinations of biomarkers that collectively offer improved discriminatory power in disease classification compared to individual biomarkers alone. In cPCA, various combinations with varying numbers of biomarkers are generated from a fixed set of input biomarkers. The number of possible combinations generated can be calculated as 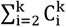.The first principal component, denoted as CP, is then extracted to represent each combination. CP serves as a comprehensive biomarker signature, representing the maximum variance direction within the biomarker combination. Finally, the performance of each CP in disease classification is evaluated and compared to that of single biomarkers.

#### Implementation Details

To systematically explore and identify potential superior components for CKD classification beyond conventional biomarkers, we applied cPCA to a set of 21 CKD-related phenotypes. Initially, we generated 2,097,130 unique combinations out of the 21 phenotypes. These combinations encompassed all possible subsets of the 21 phenotypes with varying numbers, ranging from 2 to the complete set of 21. For each combination, we extracted CP, resulting in 2 million CPs. Subsequently, we evaluated the performance of each CP in CKD classification and compared it to that of CYSC, which served as the best single marker for CKD classification.

To validate the efficacy of the identified combinations, we partitioned the dataset into a training set (70%) and a test set (30%). Notably, cPCA was exclusively performed on the training set, encompassing the 2 million combinations. The performance evaluation involved comparing the ROC curves (Receiver Operating Characteristic curves) of each CP against those of individual phenotypes. Confidence intervals for the calculated AUCs (Area Under the Curve) were computed using bootstrap methods with 2000 stratified bootstrap replicates, implemented within the R package pROC.

Combinations exhibiting significantly higher AUCs compared to CYSC were further validated using the independent test set. The final AUCs were calculated based on the entire dataset.

### Genome-wide analyses

Genome-wide association studies (GWAS) were performed by fitting linear models (for quantitative traits) or logistic models (for binary traits) implemented in PLINK2.0.^38^ All the input phenotypes were inverse-normal transformed prior to GWAS. Age, sex, and the first 20 genetic principal components were integrated into the models as covariates. SNP-based heritability and genetic correlation were estimated based on the GWAS summary statistics using linkage disequilibrium score regression (LDSC) v1.0.1^39^

## RESULTS

### Best single-markers for CKD classification

In this study, our objective was to identify novel genetic loci associated with CKD through a comprehensive multi-phenotype analysis. Prior to conducting the multi-phenotype analysis, we examined the 21 phenotypes previously linked to CKD (Table 1) in terms of their performance in classifying clinical CKD. This was evaluated by the area under the curve (AUC) of receiver operating characteristic (ROC) curves using the ICD codes for CKD as clinical outcomes (Figure 1). The biomarkers encompassed a range of physiological indicators of CKD risk, including markers of renal function, metabolic parameters, inflammation, lipid profile, and blood pressure. Notably, cystatin C (CYSC) exhibited the highest discriminatory power among the biomarkers, with an AUC range of 0.832-0.842, closely followed by estimated glomerular filtration rate (eGFR) with an AUC range of 0.825-0.835. Other biomarkers, such as blood urea nitrogen (BUN), uric acid (UA), and glycated hemoglobin (HbA1c), demonstrated moderate discriminatory performance, with AUCs ranging from 0.658 to 0.742. Conversely, other biomarkers such as vitamin D (VITD), calcium (CALC), and diastolic blood pressure (DBP) exhibited low AUC values, ranging from 0.489 to 0.525.

**Figure 1.**
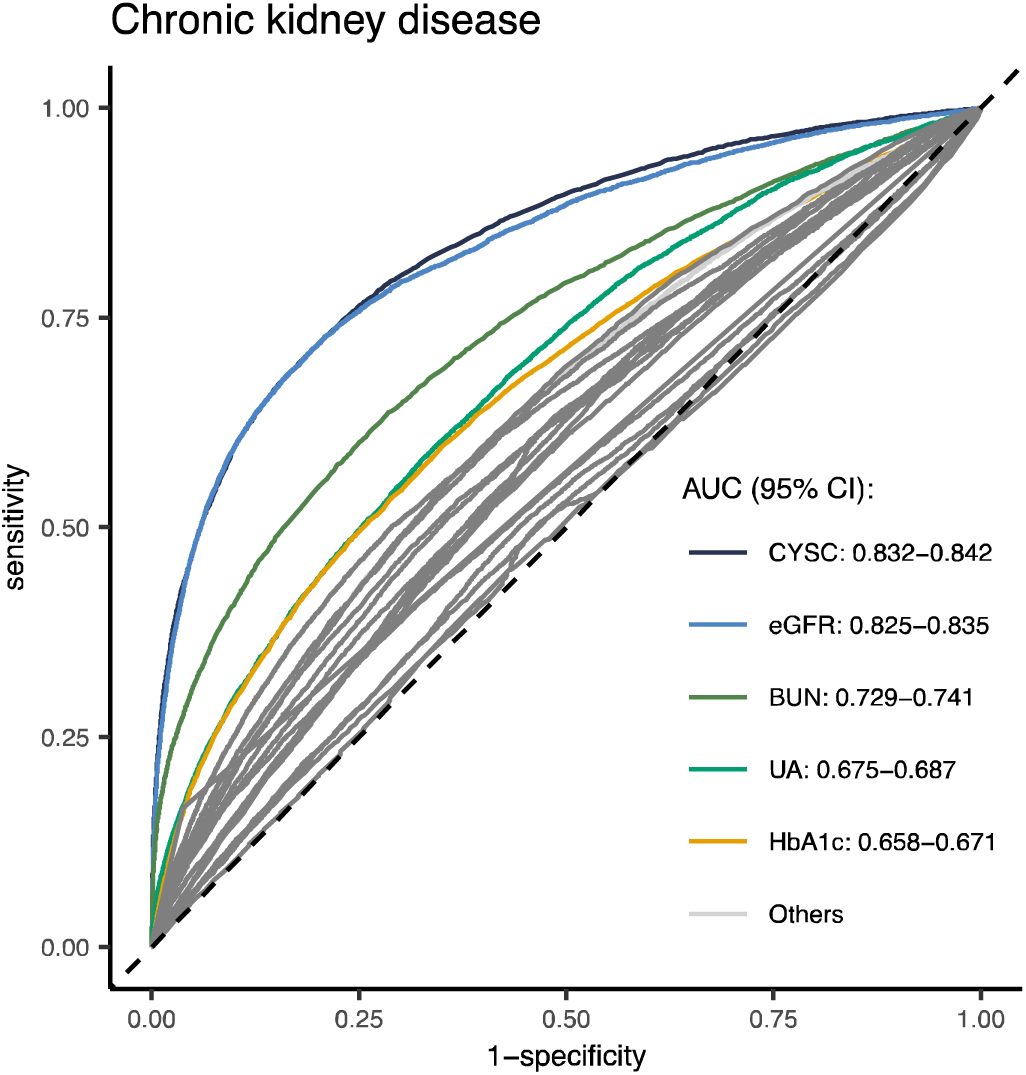
ROC curves for CKD classification of the 21 CKD-related phenotypes.

### Composite phenotypes better than single markers in CKD classification

In this multi-phenotype analysis, we developed and applied a method called combinatorial PCA (cPCA) to identify combinations of biomarkers that outperformed single markers. General steps of the cPCA method are illustrated in Figure 2A. Through cPCA application, a total of 2,097,130 composite phenotypes (CPs) were extracted from all unique combinations of 21 CKD-related biomarkers, and then evaluated for performance in CKD classification. As a result, we identified 46,562 CPs with significantly better disease classification compared to CYSC (p<2.5e-8), as assessed using ROC curves and AUC. The top ten CPs with the highest performance are listed in Table 2.

**Figure 2.**
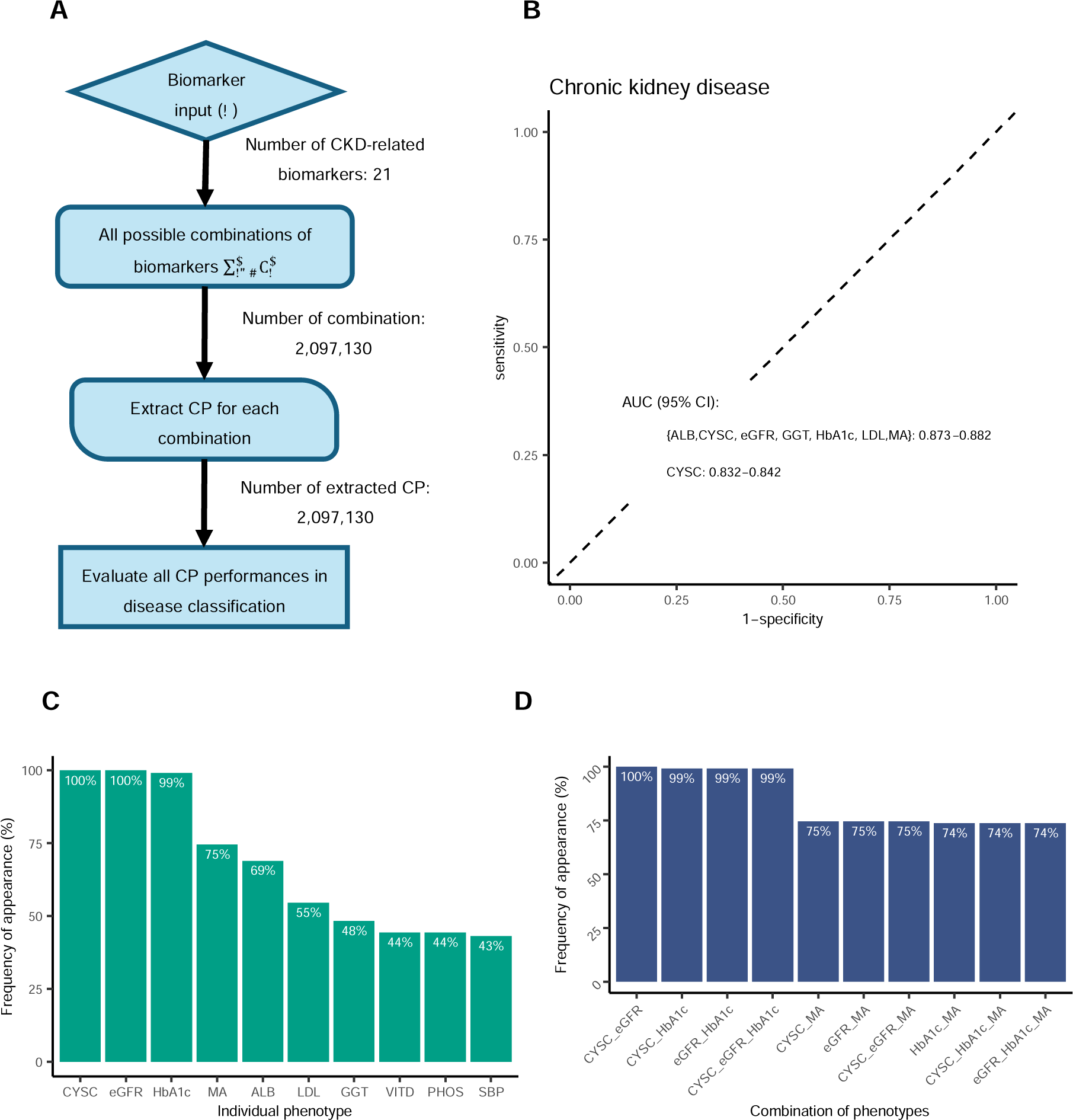
Combinatorial Principal Component Analysis (cPCA). A. Flowchart of the cPCA method. B. The ROC curve of the top CP extracted from eGFR, CYSC, MA, HbA1c, LDL, ALB, and GGT in comparison to the ROC curves of the 21 biomarkers in terms of CKD classification. C. Top 10 of the phentoypes that appeared most frequently in the 46,000 CPs. D. Top 10 of the phenoyptes pairs or triples that appeared most frequently in the 46,000 CPs.

**Table 2.**
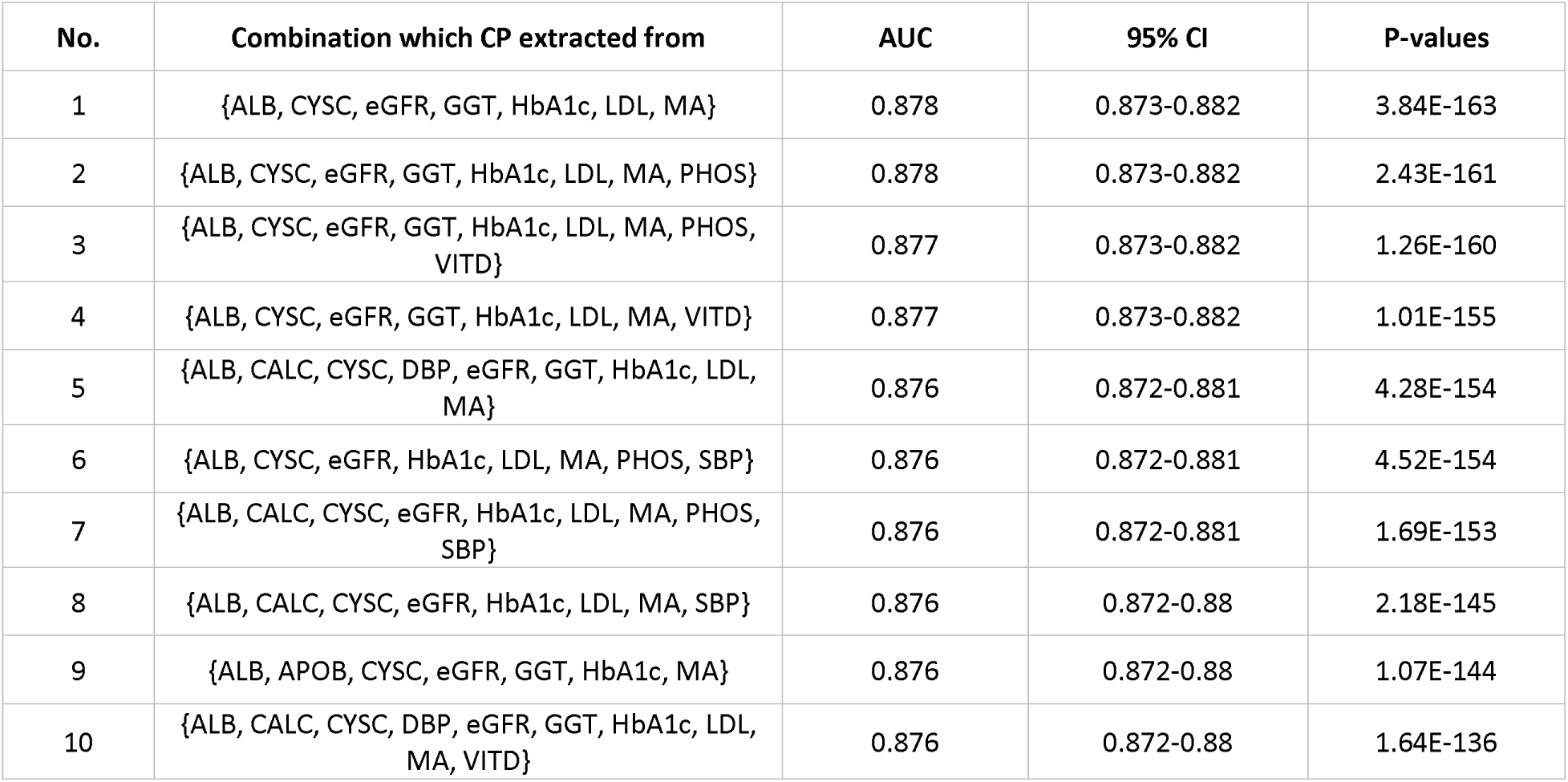
Top 10 CPs with the highest AUCs. P-values were of the tests comparing the CPs’ ROC curves to the CYSC’s ROC curve.

| No. | Combination which CP extracted from | AUC | 95% CI | P-values |
| --- | --- | --- | --- | --- |
| 1 | {ALB, CYSC, eGFR, GGT, HbA1c, LDL, MA} | 0.878 | 0.873-0.882 | 3.84E-163 |
| 2 | {ALB, CYSC, eGFR, GGT, HbA1c, LDL, MA, PHOS} | 0.878 | 0.873-0.882 | 2.43E-161 |
| 3 | {ALB, CYSC, eGFR, GGT, HbA1c, LDL, MA, PHOS, VITD} | 0.877 | 0.873-0.882 | 1.26E-160 |
| 4 | {ALB, CYSC, eGFR, GGT, HbA1c, LDL, MA, VITD} | 0.877 | 0.873-0.882 | 1.01E-155 |
| 5 | {ALB, CALC, CYSC, DBP, eGFR, GGT, HbA1c, LDL, MA} | 0.876 | 0.872-0.881 | 4.28E-154 |
| 6 | {ALB, CYSC, eGFR, HbA1c, LDL, MA, PHOS, SBP} | 0.876 | 0.872-0.881 | 4.52E-154 |
| 7 | {ALB, CALC, CYSC, eGFR, HbA1c, LDL, MA, PHOS, SBP} | 0.876 | 0.872-0.881 | 1.69E-153 |
| 8 | {ALB, CALC, CYSC, eGFR, HbA1c, LDL, MA, SBP} | 0.876 | 0.872-0.88 | 2.18E-145 |
| 9 | {ALB, APOB, CYSC, eGFR, GGT, HbA1c, MA} | 0.876 | 0.872-0.88 | 1.07E-144 |
| 10 | {ALB, CALC, CYSC, DBP, eGFR, GGT, HbA1c, LDL, MA, VITD} | 0.876 | 0.872-0.88 | 1.64E-136 |

We analyzed the phenotypic components of the 46,562 CPs that exhibited statistically significantly better performance in CKD classification compared to CYSC (Figure 2B, 2C and 2D). The top ranked CP was represented by albumin (ALB), CYSC, eGFR, gamma glutamintransferase (GGT), HbA1c, low density lipoprotein (LDL), and microalbuminuria (MA) (AUC=0.878, 95%CI=0.873-0.882). Among the other combinations, CYSC and eGFR were consistently present, with HbA1c appearing in nearly all instances. Other notable phenotypes included MA, ALB, and LDL, with appearances ranging from 75% to 55% across the combinations. Regarding pairs or triples of phenotypes, as expected, the most frequent combinations included CYSC, eGFR, and HbA1c: CYSC-eGFR pairs were present in all combinations, while CYSC-HbA1c, eGFR-HbA1c, and CYSC-eGFR-HbA1c were found in 99% of combinations.

### Genetic associations of the top 1000 CPs

The cPCA analysis identified a total of 46,562 CPs with significantly higher AUCs than that of CYSC. To uncover genetic loci associated with kidney function, we conducted a genome-wide analysis of the top 1000 CPs with the highest AUCs, as well as all 21 individual phenotypes. This analysis yielded 82 loci consistently identified in the GWASs of the top 1000 CPs (p=5e-8, Figure 3). Most of these loci were also observed in the eGFR GWAS. However, seven loci – *CST3, SH2B3*, *FTO*, *SEMA3F-AS1*, *AL359852.2*, *AC128707.1*, and *AL049757.1* - were not identified in the eGFR GWAS and were instead found in GWASs of other individual phenotypes. *SH2B3* was found in 12 out of the 21 individual-phenotype GWASs, *FTO* was found in 7 and *SEMA3F-AS*1 in 5. These 7 loci represented potentially novel genetic associations with kidney function, discovered through the multi-phenotype approach.

**Figure 3.**
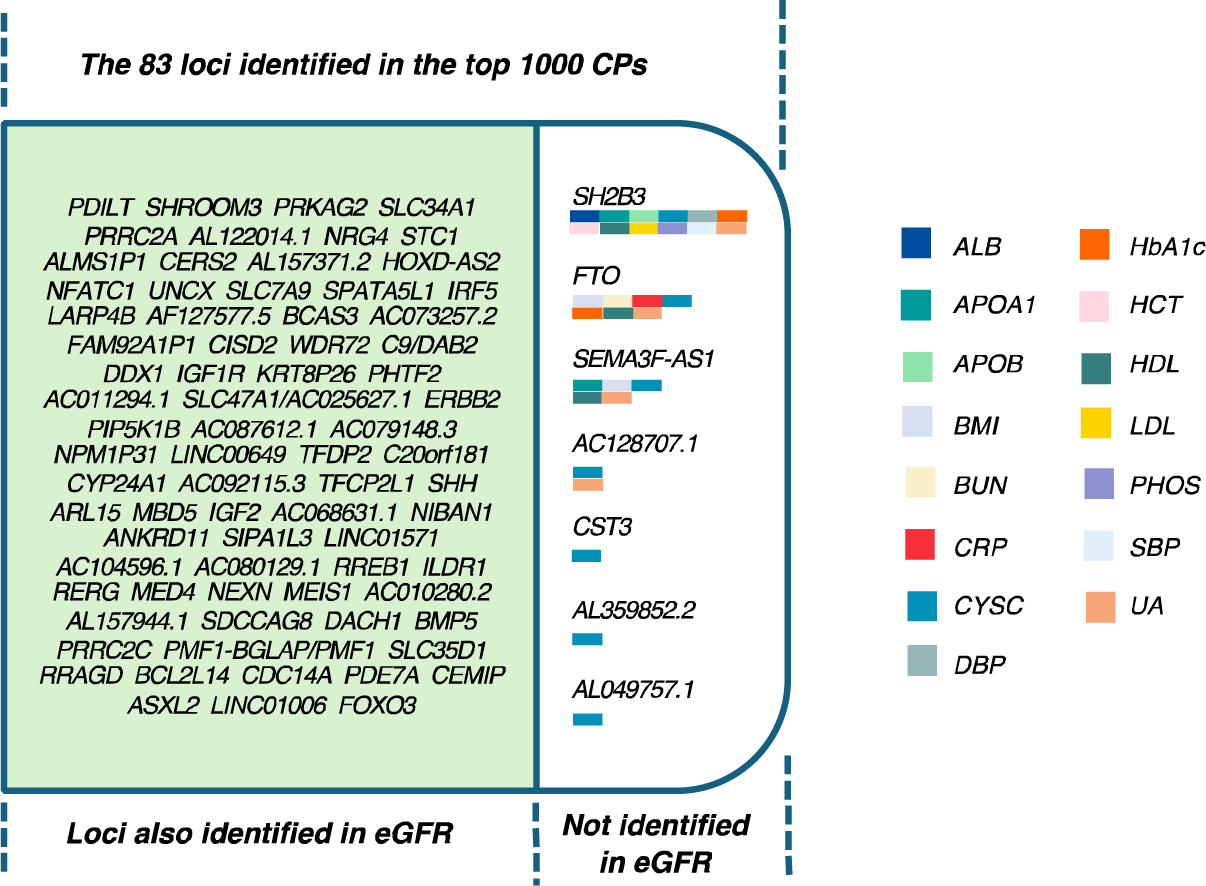
The 82 genetic loci identified in all the top 1000 CPs. Among these loci, 75 were found to overlap with those identified in the GWAS of eGFR, while 7 loci were not. Instead, these 8 loci were discovered in GWASs of other individual phenotypes, each represented by a distinct color.

### Replication analysis

We utilised the independent cohort of Irish ethnicity in the UKB dataset for our replication analysis. In the discovery group, i.e. the British cohort, the CP extracted from the combination of {eGFR, CYSC, ALB, HbA1c, GGT, LDL, and MA} was among those that had the highest AUCs for CKD classification and at the same time had the least number of phenotypes (Table 2). Therefore, we selected this combination of phenotypes to generate a new CP for the replication cohort. As a result, the new CP in the replication cohort also had significantly better performance in CKD classification compared to those of individual phenotypes (Figure 4). Out of the 7 potentially novel loci associated with kidney function, *CST3* and *SH2B3* were replicated in the Irish cohort as outlined in Table 3.

**Figure 4.**
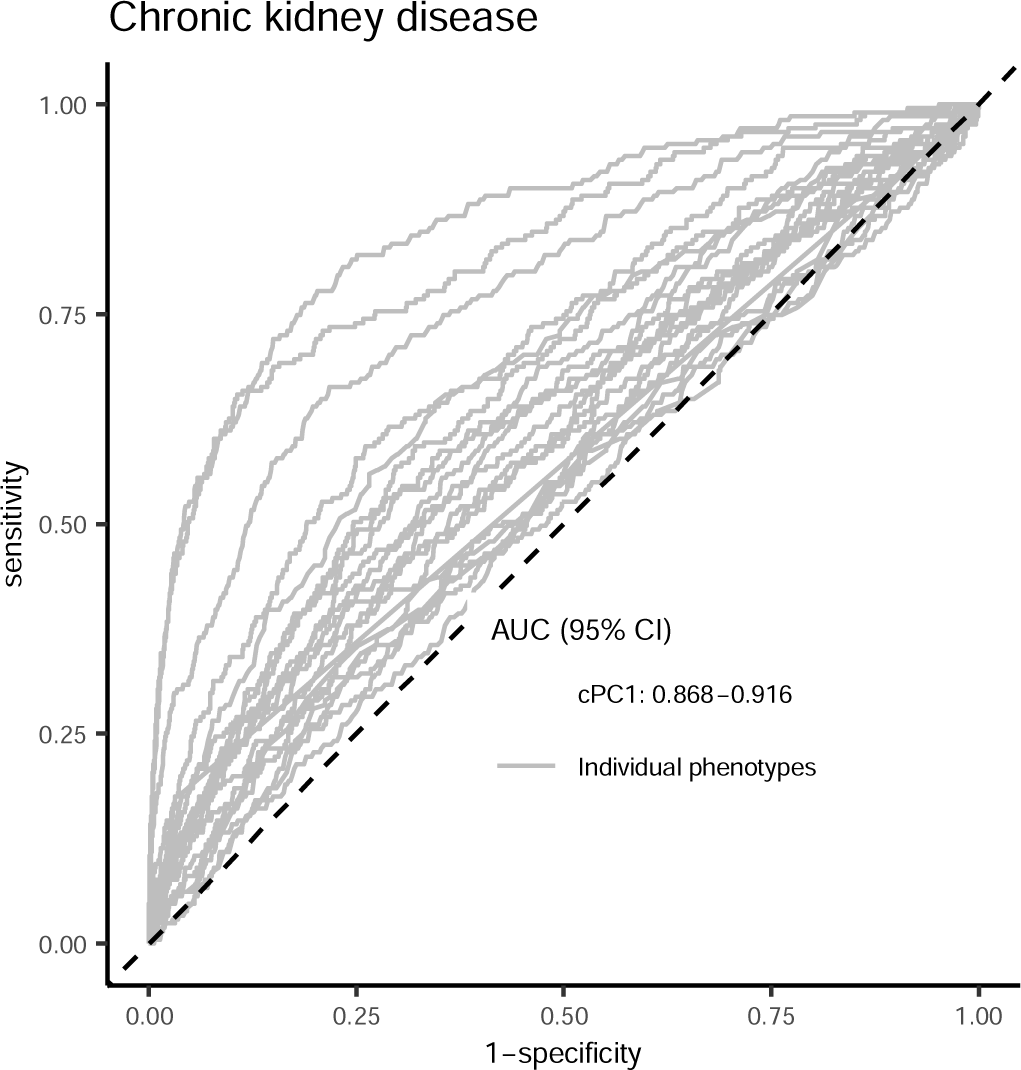
ROC curves for CKD classification of the CP and the 21 CKD-related phenotypes in the replication Irish cohort. The CP was extracted from the combinations of phenotypes {eGFR, CYSC, ALB, HbA1c, GGT, LDL, and MA}.

**Table 3.**
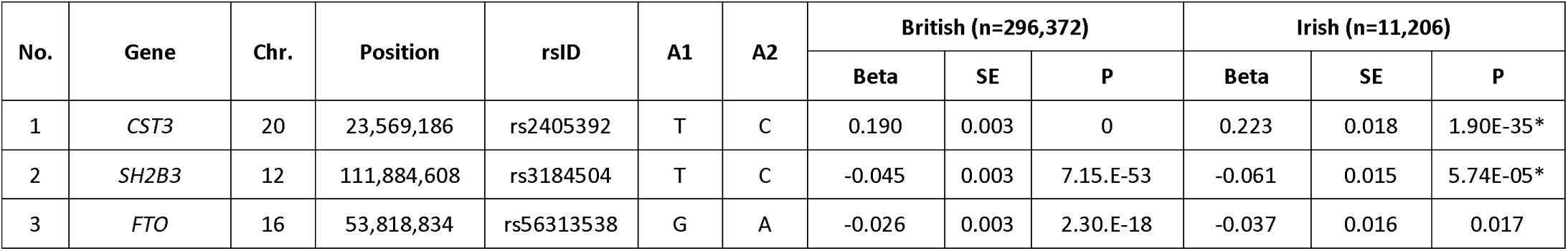
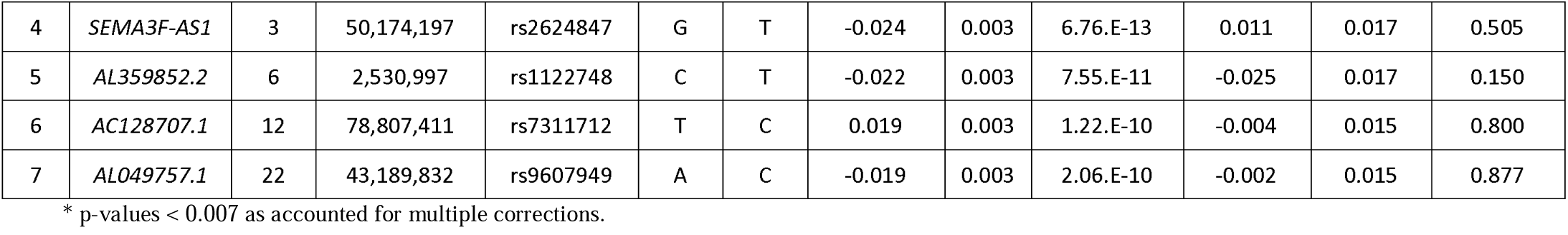
Association results of the potentially novel kidney-function loci in the discovery British group and the replication Irish group. The phenotypic outcomes were CP extracted from {eGFR, CYSC, ALB, HbA1c, GGT, LDL, and MA} for both the British group and the Irish group, respectively.

| No. | Gene | Chr. | Position | rsID | A1 | A2 | British (n=296,372) |  |  | Irish (n=11,206) |  |  |
| --- | --- | --- | --- | --- | --- | --- | --- | --- | --- | --- | --- | --- |
|  |  |  |  |  |  |  | Beta | SE | P | Beta | SE | P |
| 1 | *CST3* | 20 | 23,569,186 | rs2405392 | T | C | 0.190 | 0.003 | 0 | 0.223 | 0.018 | 1.90E-35 * |
| 2 | *SH2B3* | 12 | 111,884,608 | rs3184504 | T | C | -0.045 | 0.003 | 7.15.E-53 | -0.061 | 0.015 | 5.74E-05 * |
| 3 | *FTO* | 16 | 53,818,834 | rs56313538 | G | A | -0.026 | 0.003 | 2.30.E-18 | -0.037 | 0.016 | 0.017 |
| 4 | *SEMA3F-AS1* | 3 | 50,174,197 | rs2624847 | G | T | -0.024 | 0.003 | 6.76.E-13 | 0.011 | 0.017 | 0.505 |
| 5 | *AL359852.2* | 6 | 2,530,997 | rs1122748 | C | T | -0.022 | 0.003 | 7.55.E-11 | -0.025 | 0.017 | 0.150 |
| 6 | *AC128707.1* | 12 | 78,807,411 | rs7311712 | T | C | 0.019 | 0.003 | 1.22.E-10 | -0.004 | 0.015 | 0.800 |
| 7 | *AL049757.1* | 22 | 43,189,832 | rs9607949 | A | C | -0.019 | 0.003 | 2.06.E-10 | -0.002 | 0.015 | 0.877 |

\* p-values < 0.007 as accounted for multiple corrections.

## DISCUSSION

CKD is a common term to describe a range of diseases characterized by impaired kidney structure, or reduced kidney function over time. Because there is an incomplete understanding of the genetics for different CKD subtypes, the identification of effective drug targets has been hindered. Research has tended to focus on eGFR or other single CKD-related biomarkers, yet this approach could be inadequate for capturing the underlying CKD etiology or pathophysiology. Since CKD is associated with many individual phenotypes we reasoned that a multi-phenotype analytical approach may identify novel genetic loci relevant to CKD. Specifically we designed a combinatorial PCA algorithm (cPCA), the aim of which was to extract relevant composite phenotypes for accurate CKD classification. This involved iteratively exploring all possible combinations of the 21 input phenotypes to identify composite phenotypes that outperformed individual biomarkers in CKD classification. Over 2 million phenotypic combinations were analyzed, resulting in the identification of over 46,000 composite phenotypes with significantly higher AUCs than CYSC or any individual phenotype.

CYSC, eGFR, HbA1c, MA, ALB, LDL, and GGT were the most frequently observed phenotypes, appearing in 75% to 48% of those combinations. The frequent presence of HbA1c, ALB, LDL, and GGT alongside well-established CKD phenotypes such as CYSC, eGFR, and MA highlighted the overlap between kidney function and other aspects of human health, including blood glucose levels, cardiovascular health, and liver function.

Furthermore, we observed that although BUN and UA, which are highly correlated with eGFR, exhibited higher performance in CKD classification than HbA1c, MA, and others, they appeared less frequently in the 46,000 combinations (BUN: 22.4% and UA: 0%). This suggested that cPCA could mitigate multicollinearity by ensuring the inclusion of independent phenotypes that are not highly correlated.

Consequently, we performed GWAS of the top 1000 composite phenotypes with the highest performance in CKD classification and identified 82 loci that were consistently found in all the 1000 GWASs. As expected, most of these were also found in eGFR GWAS, including well-known CKD loci such as *UMOD/PDILT*, *SHROOM3*, and *PRKAG2*. Noteworthy was that there were 7 loci that were not identified in eGFR. They were *CST3, SH2B3*, *FTO*, *SEMA3F-AS1*, *AL359852.2*, *AC128707.1*, and *AL049757.1*. Finally, *CST3* and *SH2B3* were successfully replicated in an independent cohort. As noted, *CST3* was, in fact, already an established kidney function gene.

On the other hand, *SH2B3*, which encoded for a cytokin-signalling regulator, was less recognized for its involvement in kidney function. The index SNP rs3184504 mapped to the *SH2B3* locus is a loss-of-function variant and has been found to be associated with multiple phenotypes and diseases relating to blood pressure, blood cells, cholesterol levels, as well as cardiovascular diseases and type-1 diabetes (https://www.ebi.ac.uk/gwas/variants/rs3184504). Notably, murine animal models that were modified to be homozygous for the minor allele at rs3184504 using CRISPR-Cas9 exhibited higher blood pressure and exacerbated kidney dysfunction compared to control mice following angiotensin II infusion.^40^ This study marks the first time this missense SNP has been linked to human kidney function.

Another noteworthy finding was the identification of the *FTO* locus among the top 1000 CPs. Although the locus only reached a nominal significance level in the replication analysis, this was potentially attributed to the much lower power of replication analysis. The *FTO* gene has been associated with CKD in case-control studies,^41^. Interstingly, the identified *FTO* SNP rs17817449 in Hubacek et al^41^ was within 5.5kbp of the index SNP in our discovery GWAS and, more importantly, also reached the genome-wide significane threshold (p= 8.13E-18).

In conclusion, The cPCA method developed and applied in this study successfully identified a novel CKD locus, *SH2B3*. Moreover, this study highlighted the effectiveness of multi-phenotype approaches in uncovering novel genetic loci associated with complex diseases such as CKD, which exhibit substantial overlap with multiple other physiological components.

## DISCLOSURE

All the authors declared no competing interests.

## Data Availability

The summary statistics are publicly available in Figshare repository at https://doi.org/10.6084/m9.figshare.26122540.v1.

## ACKNOWLEDGEMENTS

This research has been conducted using the UK Biobank Resource under Application Number 60111. NKT has been supported by a QUT postgraduate scholarship. AJM has been supported by a Queensland Health Advancing Clinical Research Fellowship.

